# Cellular immune response to SARS-CoV-2 infection in humans: a systematic review

**DOI:** 10.1101/2020.08.24.20180679

**Authors:** Madhumita Shrotri, May C. I. van Schalkwyk, Nathan Post, Danielle Eddy, Catherine Huntley, David Leeman, Samuel Rigby, Sarah V. Williams, William H. Bermingham, Paul Kellam, John Maher, Adrian M. Shields, Gayatri Amirthalingam, Sharon J. Peacock, Sharif A. Ismail

## Abstract

**Introduction:** Understanding the cellular immune response to SARS-CoV-2 is critical to vaccine development, epidemiological surveillance and control strategies. This systematic review critically evaluates and synthesises the relevant peer-reviewed and pre-print literature published in recent months.

**Methods:** For this systematic review, independent keyword-structured literature searches were carried out in MEDLINE, Embase and COVID-19 Primer for studies published from 01/01/2020-26/06/2020. Papers were independently screened by two researchers, with arbitration of disagreements by a third researcher. Data were independently extracted into a pre-designed Excel template and studies critically appraised using a modified version of the MetaQAT tool, with resolution of disagreements by consensus. Findings were narratively synthesised.

**Results:** 61 articles were included. Almost all studies used observational designs, were hospital-based, and the majority had important limitations. Symptomatic adult COVID-19 cases consistently show peripheral T cell lymphopenia, which positively correlates with increased disease severity, duration of RNA positivity, and non-survival; while asymptomatic and paediatric cases display preserved counts. People with severe or critical disease generally develop more robust, virus-specific T cell responses. T cell memory and effector function has been demonstrated against multiple viral epitopes, and, cross-reactive T cell responses have been demonstrated in unexposed and uninfected adults, but the significance for protection and susceptibility, respectively, remains unclear.

**Interpretation:** A complex pattern of T cell response to SARS-CoV-2 infection has been demonstrated, but inferences regarding population level immunity are hampered by significant methodological limitations and heterogeneity between studies. In contrast to antibody responses, population-level surveillance of the cellular response is unlikely to be feasible in the near term. Focused evaluation in specific sub-groups, including vaccine recipients, should be prioritised.

## INTRODUCTION

Severe Acute Respiratory Syndrome Coronavirus 2 (SARS-CoV-2), the novel pathogen causing coronavirus disease 2019 (COVID-19), has spread globally and was declared a pandemic by the World Health Organization (WHO) on 11th March 2020.^1^ At the time of writing, there have been around 22.3m confirmed cases and 782,456 deaths reported to the WHO.^2^ Lack of pre-existing immunity to this novel and highly infectious betacoronavirus is likely to be responsible for the extraordinary surge in cases worldwide.

There has been an unparalleled global effort to characterise the immune response to SARS-CoV-2 infection, and to develop and test vaccine candidates at unprecedented speed. Understanding the patterns in individual- and population-level immunity will be key to informing future decisions on implementation of non-pharmacological interventions, broader public health policies, and strategies for vaccine delivery.

While there is a rapidly growing body of literature on the antibody response to SARS-CoV-2, much less has been published on the cellular immune response, despite its critical importance in antiviral immunity and vaccine development.

There are principally three areas of interest; firstly, the role of T cells in viral control and immunopathogenesis during acute SARS-CoV-2 infection; secondly the role of T cells in establishing durable protective immunity against reinfection; finally, the relevance of pre-existing cross-reactive cellular immunity from endemic human coronaviruses (HCoV), or SARS-CoV-1.^3^

This paper focuses on summarising current understanding of the cellular response to SARS-CoV-2 infection, specifically exploring the role that T cell-mediated immunity might play in resistance to severe infection, clinical and virological recovery, and long-term protection – while recognising the dynamic interdependence of the two arms of the adaptive immune response. It is the second of two linked papers^4^ summarising results from a wide-ranging systematic review of peer-reviewed and pre-print literature on the human adaptive immune response to SARS-CoV-2 infection.

## METHODS

A systematic review was carried out according to the Preferred Reporting Items for Systematic Reviews and Meta-Analyses (PRISMA) guidelines. The protocol was pre-registered with PROSPERO (CRD42020192528).

### Identification of studies

Keyword-structured searches were performed in MEDLINE, Embase, COVID-19 Primer and the Public Health England library^5^ for articles published between 01/01/2020-26/06/2020. A sample search strategy can be found in **Supplementary Appendix A**. We also consulted subject area experts to identify relevant papers not captured through the database searches.

### Definitions, inclusion and exclusion criteria

We included studies in all human and animal populations, and carried out in all settings (laboratory, community and clinical - encompassing primary, secondary and tertiary care centres), relevant to our research questions. We excluded case reports, commentaries, correspondence pieces or letter responses, consensus statements or guidelines and study protocols. We included studies reporting on any aspect of the T cell response, irrespective of follow-up duration, and on correlates of that response. We defined “correlates” to include (among others) age; gender; ethnicity; the presence of intercurrent or co-morbid disease e.g. diabetes, cardiovascular, chronic respiratory disease; and primary illness severity, proxied by the WHO’s distinction between “mild”, “moderate”, “severe” and “critical” COVID-19,^6^ or by requirement for intensive care.

### Selection of studies

Studies were independently screened on title, abstract and full text by two team members (working across four pairs), and disagreements arbitrated by one of the review leads.

### Data extraction, assessment of study quality, and data synthesis

Data were extracted in duplicate from each included study into a dedicated Microsoft Excel template (**Supplementary Appendix B**). Pre-prints of subsequently published peer reviewed papers were included, and results extracted where substantial differences in reported data were identified; if little difference was found, only the peer-reviewed version was retained.

Critical appraisal for each included study was performed in duplicate using a version of the MetaQAT 1.0 tool that was adapted for improved applicability to the basic science and laboratory-based studies that are common in this field.^7^ The adapted MetaQAT tool was used to gather both qualitative feedback on study quality, and scaled responses (yes/no/unclear) to questions around study reliability, internal and external validity, and applicability, in turn converted into weighted scores for each paper. Accordingly, studies were assigned a “high”, “medium” or “low” quality grading. Full details of this process can be found in **Supplementary Appendix C**.

Due to the degree of methodological heterogeneity across included studies, formal meta-analysis was not performed. Results are synthesised narratively in the sections that follow.

## RESULTS

### Descriptive overview of included studies

A total of 9,223 records were identified through searches conducted for the review after de-duplication, and a further five through expert consultation, of which 61 papers were included (see PRISMA flowchart in **figure 1**).

**Figure 1.**
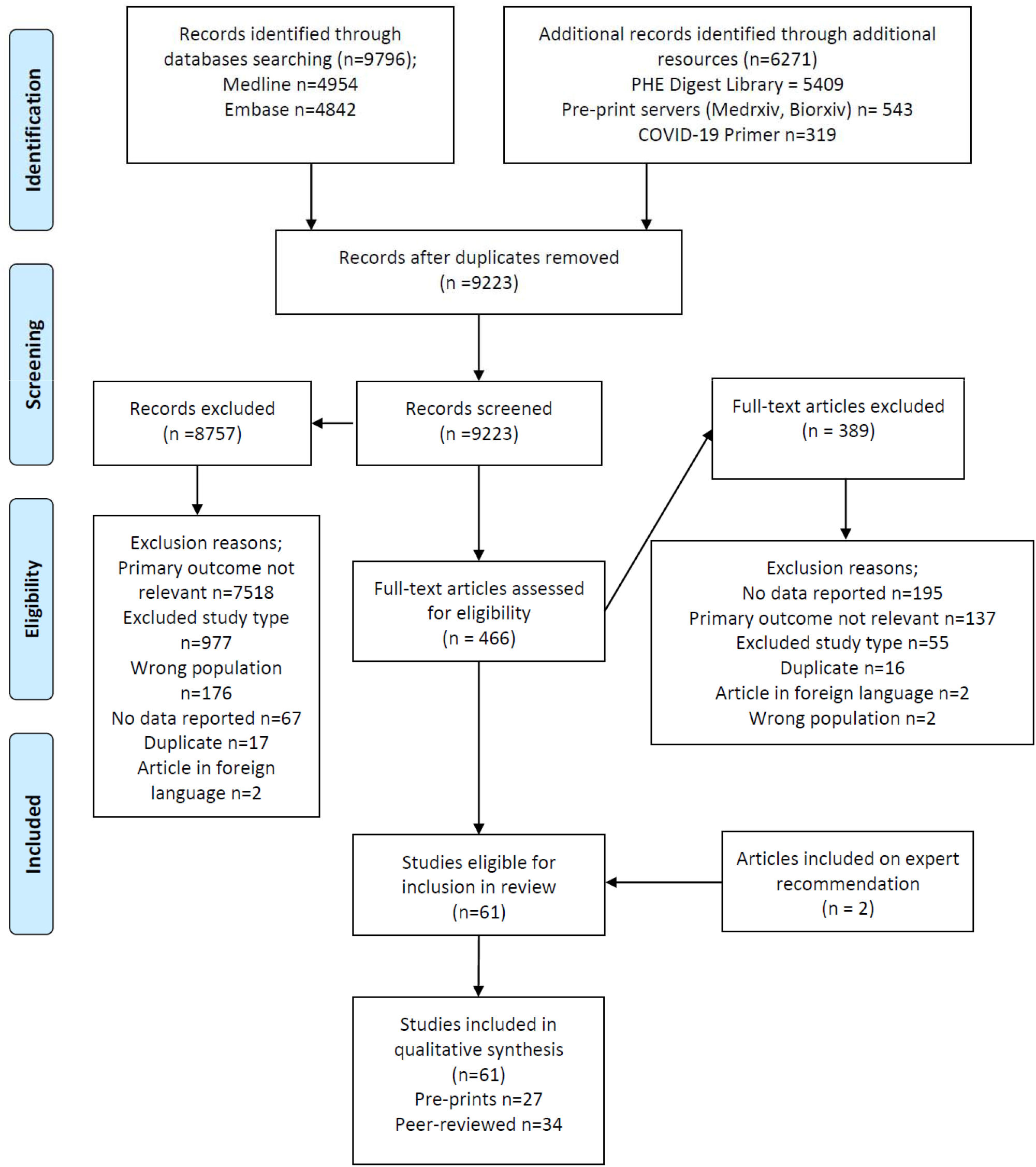
PRISMA flowchart documenting the search and screening process for this review.

Key characteristics of included studies are further summarised in **table 1**. Of the included reports, 34 (58%) were peer-reviewed journal papers. The most common designs were case-control (n=26, 43%) and cohort (n=22, 36%), with 50 studies (82%) considering human participants sampled from hospital settings, and most originating from China (n=32, 52%). Only five studies (8%) specifically examined cellular responses in children; while only one study examined differences by gender, and none by ethnicity (see **table 2**). Most studies were rated of medium quality (n-44, 72%), with ten (16%) achieving a high-quality rating.

**Table 1.**
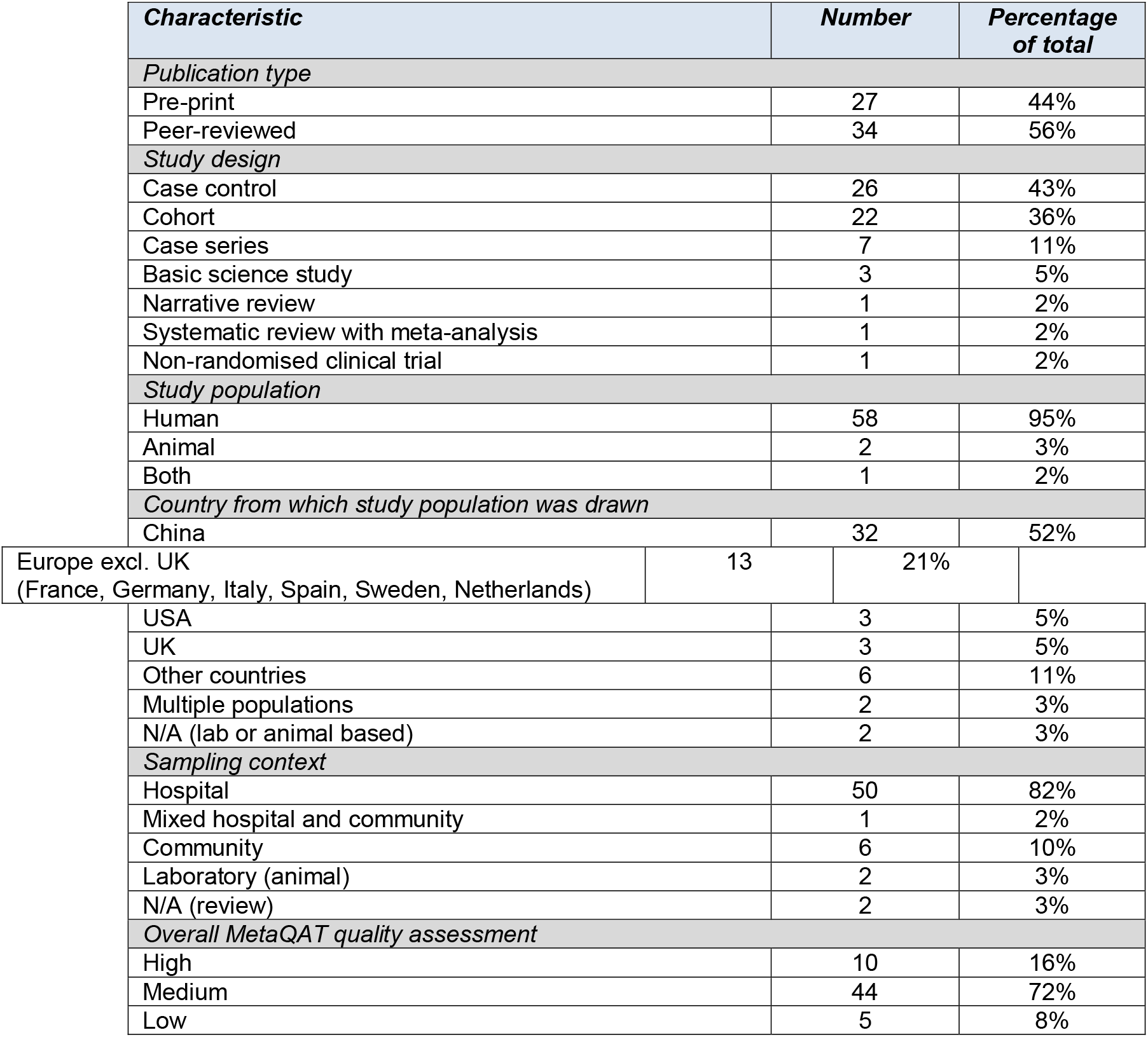
Summary of descriptive statistics for included studies.

| **Characteristic** | **Number** | **Percentage of total** |
| --- | --- | --- |
| **Publication type** |  |  |
| Pre-print | 27 | 44% |
| Peer-reviewed | 34 | 56% |
| **Study design** |  |  |
| Case control | 26 | 43% |
| Cohort | 22 | 36% |
| Case series | 7 | 11% |
| Basic science study | 3 | 5% |
| Narrative review | 1 | 2% |
| Systematic review with meta-analysis | 1 | 2% |
| Non-randomised clinical trial | 1 | 2% |
| **Study population** |  |  |
| Human | 58 | 95% |
| Animal | 2 | 3% |
| Both | 1 | 2% |
| **Country from which study population was drawn** |  |  |
| China | 32 | 52% |
| Europe excl. UK<br>(France, Germany, Italy, Spain, Sweden, Netherlands) | 13 | 21% |
| USA | 3 | 5% |
| UK | 3 | 5% |
| Other countries | 6 | 11% |
| Multiple populations | 2 | 3% |
| N/A (lab or animal based) | 2 | 3% |
| **Sampling context** |  |  |
| Hospital | 50 | 82% |
| Mixed hospital and community | 1 | 2% |
| Community | 6 | 10% |
| Laboratory (animal) | 2 | 3% |
| N/A (review) | 2 | 3% |
| **Overall MetaQAT quality assessment** |  |  |
| High | 10 | 16% |
| Medium | 44 | 72% |
| Low | 5 | 8% |

**Table 2.** Evidence on clinical and demographic correlates of cellular response to SARS-CoV-2 infection from studies included in this review [* Disease severity was defined in various ways in included studies; for some according to intensive care unit admission; a number used the Chinese National Health Commission definition (The Notice of Launching Guideline on Diagnosis and Treatment of the Novel Coronavirus Pneumonia (NCP). 5th ed. Available online at: http://www.nhc.gov.cn/yzygj/s7653p/202002/3b09b894ac9b4204a79db5b8912d4440/files/7260301a393845fc87fcf6dd52965ecb.pdf (accessed February 18, 2020))]

| Category | Correlate | Dimension or sub-population | Findings |
| --- | --- | --- | --- |
| Clinical | Disease severity* | Asymptomatic or pauci-symptomatic | One study evaluated T cell responses in asymptomatic patients (n=20) and found little change in the circulating T cell frequencies within this group.<sup>12</sup> |
|  |  | Moderate disease | Reduced numbers of both CD4<sup>+</sup> and CD8<sup>+</sup> T cells in moderate and severe cases, alongside increased numbers of activated CD4<sup>+</sup> and CD8<sup>+</sup> T cells expressing PD-1 or Tim-3; as well as potential reductions in cytotoxic potential and polyfunctionality reported in one narrative review.<sup>3</sup> |
|  |  | Severe or critical disease | *Cell counts*<br>A medium quality meta-analysis found that patients with severe disease had statistically significant, two-fold decreases in both CD4<sup>+</sup> and CD8<sup>+</sup> T cells, as well as in CD3<sup>+</sup> T cells (1.7-fold) and overall lymphocyte number (1.44-fold), alongside statistically significant increases in neutrophils (1.33-fold) and overall leukocytes (1.2-fold).<sup>16</sup><br>A large study (N=599) by Diao et al. reported reduced total, CD4<sup>+</sup>, and CD8<sup>+</sup> T cells being associated with more severe disease, comparing n=43 ICU-admitted patients with non-ICU-admitted patients, and comparing critical/severe non-ICU patients with mild/moderate non-ICU patients (as per Chinese national definitions*).<sup>13</sup><br>Other large studies<sup>10,32,34,59</sup> showed comparable findings, and 3 studies also reported reduced CD3<sup>+</sup> cells in more severe disease;<sup>10,34,59</sup> however, Liu et al. only found significant cell count differences for critical vs severe disease, and not for severe vs moderate disease.<sup>59</sup><br>*Cell ratios*<br>Six studies reported marked increases in CD4/CD8 ratio (due to increases in CD4<sup>+</sup> but reductions in CD8<sup>+</sup> cells) in severe and critical patients compared to those with moderate disease.<sup>9,22,30,34,60</sup> The last of these also showed CD8<sup>+</sup> T cell counts were much slower to normalise than CD4<sup>+</sup> in patients with severe disease.<sup>60</sup><br>Two studies however, reported significant reductions in CD4<sup>+</sup>, but not in CD8<sup>+</sup>, T cells in severe disease (n=452), or 'aggravated' disease, defined as clinically progressive at 7 days (n=17).<sup>61,62</sup><br>A small study from Iran reported increased CD8 expression in ICU patients relative to healthy controls, quantified by flow cytometry as mean fluorescence intensity (MFI), with no significant differences seen in CD4/CD8 ratio, or CD4<sup>+</sup> T cell MFI.<sup>63</sup> |
|  | Clinical endpoint | Survival vs non-survival | Two studies with large cohorts followed up COVID-19 patients until death or discharge, both conducting multivariate analysis. Luo et al. (n=1018), reported significantly lower CD3<sup>+</sup>, CD4<sup>+</sup> and especially CD8<sup>+</sup> counts in non-survivors than survivors, and found that CD8<sup>+</sup> T cell counts <165 cells/μL (OR 5.93) were independently associated with mortality after adjustment for age, sex and comorbidities.<sup>36</sup> Liu et al (n=340) reported that lower helper T cells (OR 0.22) and higher CD4/CD8 ratio (OR 4.8) were highly significant predictors of mortality.<sup>64</sup><br>Whilst also reporting lower CD8<sup>+</sup> counts in non-survivors throughout the disease course, Wang et al (n=157) also found that non-survivors had lower CD4<sup>+</sup> counts only evident in middle and late stages of disease, and that non-survivors had a lower CD4/CD8 ratio.<sup>65</sup><br>Based on 28 deaths amongst 187 patients, Xu et al demonstrated that total T cell counts <500/μl, CD3<sup>+</sup> counts <200/μl, CD4<sup>+</sup> or CD8<sup>+</sup> counts <100/μ as well as B cell counts <50/μL, were significantly associated with risk of in-hospital death, however this is only on univariate analysis.<sup>10</sup> |
|  |  |  | In a cohort of n=548, Chen et al reported significantly elevated neutrophil-to-lymphocyte ratio (NLR), platelets-to-lymphocytes ratio (PLR), reduced peripheral CD3<sup>+</sup>, CD4<sup>+</sup> and particularly CD8<sup>+</sup> counts in non-survivors.<sup>34</sup> He et al. (n=204) reported that T cell levels continued to fall until death in non-survivors, whilst in survivors with severe disease, levels increased after 15 days and normalised after 25 days of treatment.<sup>32</sup> |
|  |  | RNA persistence | Four small but high or medium quality clinical cohort studies from China showed that slower resolution of PCR-positivity is associated with reductions in peripheral T cells.<br>Jiang et al. (n=23) found that the baseline abnormalities in CD3<sup>+</sup>, CD4<sup>+</sup> and CD8<sup>+</sup> T cells underwent robust recovery in patients who became RNA negative 2 weeks after diagnosis, whilst they did not do so in those who remained persistently positive.<sup>14</sup><br>Liu et al compared 37 cases who remained positive at day 20, with 37 patients at their point of diagnosis, as well as 54 healthy controls, and showed that both the persistently positive and control groups had higher CD3<sup>+</sup> and CD4<sup>+</sup> levels, suggesting that these subsets do normalise despite viral persistence.<sup>40</sup><br>In a similar study, though with a persistence threshold of 15 days, Dong et al (n=18) also found global reductions across CD3<sup>+</sup>, CD4<sup>+</sup> and CD8<sup>+</sup> subsets for persistent positives, which increased between admission and discharge; as well as significant negative correlation between overall T cell count and duration of positive nucleic acid test.<sup>66</sup><br>Liu et al. (n=39) also reported higher global T and B cells in patients becoming RT-PCR negative within 14 days.<sup>67</sup> |
|  | Co-morbid | disease status | Three studies considered the effect of comorbid status, all originating from China and spanning patients with non-severe, severe and critical clinical presentations.<sup>32,67,68</sup> Two had significant methodological limitations.<sup>67,68</sup><br>One study (n=204) found significantly lower total lymphocyte and lymphocyte subset counts in patients with comorbidities compared with those without (though “comorbidities” not defined).<sup>32</sup><br>The second (n=39) found statistically significant differences in CD8<sup>+</sup> counts between patients with comorbid disease and those without (p=0.046), but no difference in CD4<sup>+</sup> counts - although here again the range of comorbidities considered was not defined.<sup>67</sup><br>The final study compared outcomes in a paediatric cohort with or without “allergic disease” (not clearly defined) and showed no effect on clinical course, total lymphocyte or lymphocyte subset counts.<sup>68</sup> |
| Demographic | Age | Older adults | A high-quality clinical cohort study and a medium-quality case-control study, both from China, reported lower T cell total and subset counts, including CD3<sup>+</sup>, CD4<sup>+</sup>, CD8<sup>+</sup> subsets, for older patients aged 60 or over.<sup>13,32</sup> |
|  |  | Children | Four medium-quality studies - 1 case control and 3 case series - considered cellular responses in children in samples from China, all showing comparable CD3<sup>+</sup>, CD4<sup>+</sup> and CD8<sup>+</sup> counts to healthy paediatric controls, or where the comparison group was adults, higher T cell counts across subsets.<sup>21,69–71</sup> However, potential confounders such as disease severity or comorbidities were not controlled for in these studies. |
|  | Sex |  | One medium-quality case series (n=27) from China examined differences in cytokine secretion by sex of cases, showing reductions in CD4<sup>+</sup> and CD8<sup>+</sup> count for all patients irrespective of gender but more generalised cytokine responses were observed among male participants than females, for IL-6, TNF-α and procalcitonin – although the statistical significance of these differences was not tested.<sup>72</sup> |

### Acute phase T cell response and association with cytokine release syndrome

#### General features of the T cell response in the acute phase

The majority of included papers commented on general aspects of the cellular response to SARS-CoV-2 infection in the acute phase of illness, though the duration of this period was not explicitly defined. Methods used to quantify the T cell response varied between studies; for example, Laing et al partnered a total lymphocyte count from a full blood count and flow cytometry to derive estimates of absolute T cell subset counts based on the gated percentages, while other studies used direct quantification of lymphocyte subsets, such as TruCount™ and Flow-Count™ Fluorosphere technology.

Higher quality studies consistently found evidence for reduction of total peripheral T cell counts in symptomatic adult patients during the acute phase, often accompanied by increased activation of remaining T cells and evidence of functional ‘exhaustion’, as defined by expression of the markers PD-1 and Tim-3; however findings regarding specific subsets were more mixed. Three well-designed cohort studies^8–10^ showed reductions in both CD4^+^ and CD8^+^ T cell counts in clinical cohorts ranging in size from 30 to 187 patients, while two found evidence of greater reductions in CD8^+^ (cytotoxic) than CD4^+^ (helper) T cells.^8,9^ A cohort study (n=17 patients) only found evidence of reduction in CD4^+^ but not CD8^+^ T cell counts on comparing patients with ‘aggravated’ (or clinically progressive) with non-aggravated disease.^11^ A cohort study of 64 patients from Italy showed that T cell frequencies were maintained in patients with mild and asymptomatic disease.^12^ Broadly similar findings emerge from a range of high-quality case-control studies, typically with much larger sample sizes. Three hospital-based case-control studies with sample sizes ranging from 102 to 522 patients found evidence of globally reduced lymphocyte counts (CD3^+^, CD4^+^ and CD8^+^ T cells) in the acute phase.^13–15^ These findings were also reflected in two summary reviews.^3,16^ The first, a medium-quality meta-analysis incorporating data on 5,912 patients across 35 published/pre-print reports, showed that total numbers of B cells, T cells and natural killer (NK) cells were all significantly decreased in COVID-19 patients’ peripheral blood.^16^ This picture of peripheral T cell lymphopenia in COVID-19 patients is reinforced by findings from a larger body of medium- to low-quality observational studies.e.g.16–18 Notably, four studies considering cellular responses in paediatric COVID-19 cases universally demonstrated comparable T cell counts to healthy paediatric controls, or higher counts when compared against adult cases.^19–21^ The one study to evaluate responses in asymptomatic adult cases (n=20) found little change in the circulating T cell counts within this group also.^12^

Five studies provided more detailed analysis of T cell phenotypes in severe and/or critical disease.^12,22–25^ A high-quality study by Anft et al. (n=53) found significant peripheral depletion in critical patients of activated (e.g. HLA-DR^+^) memory/effector T cells that co-express tissue migratory markers (e.g. CD11a), when compared to severe and moderate cohorts.^22^ Lower frequencies of terminally differentiated T-cell subsets (TEMRA) were found in patients with both severe and critical disease. Importantly, recovery from acute respiratory distress syndrome (ARDS) was accompanied by a restoration of CD11a^+^ T cell subsets. Two studies of critically ill patients identified stronger inflammatory cytokine T cell responses to spike protein,^22^ and to spike, membrane and nucleocapsid proteins, with greater reactivity by CD4^+^ compared to CD8^+^ cells ^23^ within this group, respectively. Carsetti et al. reported an overall increase in activated (e.g. HLA-DR^+^) CD4^+^ T cells in 16 patients across both mild and severe disease but found that HLA-DR^+^ CD8^+^ cells were specifically increased in severe disease.^12^ Two studies also found increased numbers of activated T cells in patients with severe and critical disease, with reversal upon disease remission.^24,25^

Accompanying T cell dysregulation, a cytokine release syndrome (CRS)-like clinical picture occurs in many patients with severe SARS-CoV-2 infection.^26^ Elevated levels of many pro-inflammatory cytokines, such as IL-6, and to lesser degree, IL-10, and TNF-α were identified in patients in four studies.^3,27–29^ Concentrations of pro-inflammatory cytokines such as IL-6 positively correlated to severity of disease and with lymphopenia.^9,13,16,17,24,30–36^ A large peer-reviewed study with 1,018 participants reported over ten-fold increases in IL-6 levels amongst COVID-19 cases, and found that serum IL-6 >20pg/mL was strongly associated with in-hospital mortality (OR 9.78, p<0.001) on an adjusted multivariable regression analysis.^36^ A pre-print systematic review reported 1.93-fold increases in IL-6 and 1.55-fold increases in IL-10 for severe patients.^16^ In line with this, another large study (n=548) reported significantly increased IL-6 levels in non-survivors compared with survivors.^34^ Correspondingly, levels of IL-6 and IL-10 appeared to be negatively correlated with total T cell and subset counts across all included studies, and showed normalisation in tandem with clinical resolution.^13^ Findings for IL-1, IL-2, IL-4 and IL-8 were more mixed.^13,16,24,32,33,35,37^

#### Dynamics of the T cell response over time during the acute phase

Seven studies reported longitudinal data on the cellular response, mostly focusing on within-hospital trends, with a maximum follow-up range of 14-44 days following symptom onset.^10,14,17,32,38–40^ Two large high-quality case-control studies (n=103 and n=187) found that low T cell counts on admission increased steadily over the course of admission. Subsequent recovery of lymphocyte count was roughly consistent with clinical improvement.^10,14^ One study found evidence of significant decreases in counts of CD3^+^ T, CD4^+^ T, CD8^+^ T, and NK cells in COVID-19 patients compared with healthy controls (all p<0.05) on admission. In a subset of n=23 patients followed up two weeks after initial presentation, those with newly negative PCR results showed the most dramatic recoveries in T cell subset counts.^14^ Two studies reported longitudinal trends in detail at regular follow-up intervals; the first, a cohort study from Italy involving 18 patients (nine mild and nine severe cases), found that low total lymphocyte counts in severe cases were stably maintained for up to 20 days post-admission, with little discernible difference between T cell subsets.^17^ The second, a French cohort study (n=15) of predominantly elderly patients admitted to intensive care, found that CD8^+^ counts fell to their lowest value by days 11-14 after symptom onset (p=0.03), with recovery thereafter, but noted a slightly later nadir for CD4^+^ (days 19-23) and with no significant change in the overall CD4/CD8 ratio throughout the 35-day follow-up period.^39^

### Correlates of the T cell response

The number of studies addressing demographic and clinical correlates of the cellular response was small and many potentially important variables such as ethnicity were not addressed. Key findings from this literature are summarised in **table 2**. The largest single body of work examined relationships between T cell response and disease severity, based predominantly on studies in the hospital setting. Definitions of clinical severity employed in these studies were variable (most as per WHO, however some were based on Chinese national guidance).

### Viral cross-reactivity of T cells

Eight studies explored cross-reactivity of T cells between SARS-CoV-2 and related human coronaviruses within small adult-only cases and controls.^41–48^. Using activation-induced marker assays, Grifoni et al. detected SARS-CoV-2-reactive CD4^+^ T cells against a range of spike and non-spike epitopes in 12/20 pre-pandemic US donors^42^ while Weiskopf et al reported low levels of cross-reactivity in only 2/10 pre-pandemic German donors.^48^ Using IFN-γ ELISpot, Gallais et al found some T cell cross-reactivity mainly to the S2-domain in 5/10 pre-pandemic French donors^41^ and Le Bert et al found T cells specific to nucleocapsid protein (NP) and non-structural proteins 7 and 13 (NSP7, NSP13) in SARS-CoV-1/2 unexposed donors.^43^ The latter Singapore-based study also reported robust SARS-CoV-2 NP-reactivity in T cells from SARS-CoV-1 convalescents, with these memory cells persisting for 17 years after the SARS outbreak.^43^

Amongst controls recruited during the pandemic, but confirmed as antibody- and PCR-negative, spike-reactive T cells were demonstrated in 23/68 controls in a high-quality German study;^44^ and in 12/14 controls in a medium-quality Russian study, including one household contact of a COVID-19 case. The latter study also included a smaller group of pre-pandemic donors (n=10), who had significantly lower frequency and magnitude of reactivity than the controls recruited during the pandemic, hinting at a possible protective effect of cross-reactive T cells.^45^ In contrast, Peng et al. found no SARS-CoV-2-specific T cell responses in either pre-pandemic or during-pandemic antibody-negative UK controls (n=19).^46^

Notably, studies consistently found a lower frequency and magnitude of T cell response as well as a differential pattern of immunodominance in reactive unexposed controls relative to SARS-CoV-2 convalescents, with low homology between COVID-19 convalescent T cell epitopes and known epitopes from other HCoV. Interestingly, an Australian study found that frequencies of T follicular helper (TFH) cells specific to HCoV-HKU1 were higher amongst COVID-19 convalescents (n=41) than uninfected controls (n=27), suggesting boosting of HKU1-specific responses following SARS-CoV-2 exposure, and hinting at a coronavirus-specific TFH response.^47^

### Characterisation of T-cell subpopulations and protective immunity

Twelve studies characterised T-cell subpopulations, including magnitude, functionality and phenotypic characteristics, post-acute COVID-19 infection. Timing of sampling differed both within and between studies (**supplementary appendix D**). One French contact-tracing study demonstrated SARS-CoV-2-specific T cell responses against structural (spike, membrane, nucleocapsid) and accessory proteins in all nine index cases, in samples collected at 47-69 days post symptom-onset, as well as in 6/8 PCR-negative or untested contacts (of whom five were symptomatic), in samples collected up to 80 days post-onset.^41^ A UK-based study of COVID-19 convalescents (28 mild cases, 14 severe cases) characterised the T cell response using IFN-γ ELISpot assays on samples taken at least 28 days post symptom onset.^46^ A strong and broad SARS-CoV-2-specific T cell response was generally elicited but varied between individuals. T cell response breadth (p=0.010) and magnitude (p=0.002) were significantly higher in patients who recovered from severe disease in comparison to mild cases. Sub-set evaluation demonstrated CD8^+^ T cells mediated a greater proportion of responses detected to spike and membrane (M) or NP epitopes. No difference in the levels of polyfunctional T cells was observed between mild and severe disease. Differences were observed in the cytokine profiles of CD8^+^ T cells targeting different viral antigens, with the M/NP-specific CD8^+^ T cells displaying wider functionality compared to those targeting spike protein (p=0.0231). In those with mild disease, M/NP-specific CD8^+^ T cells were significantly higher than spike-specific T cells. This trend was not observed in those with severe disease.^46^

These findings complement the study by Grifoni et al (discussed above) which found that NP, M and spike contain the immunodominant epitopes for both CD4^+^ and CD8^+^ T cells.^42^ No significant differences in the cytotoxic potential was detected between mild and severe disease. Specific SARS-CoV-2-reactive T cells were not frequently observed in healthy, unexposed individuals. Furthermore, the magnitude of T cell responses in COVID-19 patients correlated with related antibody titres (anti-spike, anti-RBD and anti-NP). Another study stimulated peripheral blood mononuclear cells (PBMCs) from 18 COVID-19 patients ranging in disease severity with two overlapping peptide pools spanning the full spike region.^44^ Twelve patients had detectable CD4^+^ T cell reactivity against the first peptide pool, which contained N-terminal epitopes including the RBD. Fifteen patients displayed reactive CD4^+^ T cells against the second peptide pool, which contained C-terminal epitopes processing higher homology with HCoVs. Among the non-reactive cases most had critical disease.^44^

Le Bert et al assayed peripheral blood T cell responses to NP and NSP7 and NSP13 of the large SARS-CoV-2 proteome using an IFN-γ ELISPOT assay. Samples were obtained from 24 individuals who had experienced mild to severe COVID-19. For all patients, IFN-γ spots were observed following stimulation with NP peptide and nearly all displayed responses against multiple regions of NP. A further sub-analysis demonstrated T cell recognition of multiple regions of SARS-CoV-2 NP among recovered patients (8/9).^43^

Six studies reported on the phenotypic and target profile of T cell subsets. One study performed an in-depth characterisation of humoral and cellular immunity against the spike protein in samples taken from 41 adults who had recovered from mild-moderate SARS-CoV-2 infection (five requiring hospitalisation but not mechanical ventilation) and 27 controls. Expanded populations of spike-specific memory B cells and circulating (c)TFH cells (which play a critical role in supporting antigen-specific B cells to initiate and maintain humoral immune responses) were detected.^47^ The frequencies of unstimulated cTFH cells were comparable between SARS-CoV-2 convalescent and uninfected groups. In general, robust cTFH cells activity to the SARS-CoV-2 spike protein was observed among the convalescent group, whereas responses to RBD-specific cTFH were significantly lower (p<0.0001). The antigen reactivity of spike-specific non-cTFH CD4 memory (CD3^+^CD4^+^CD45RA^-^CXCR5^-^) cells revealed similar trends with strong recognition of SARS-CoV-2 and smaller frequencies of RBD-specific T cells. High plasma neutralisation activity was also found to be associated with increased spike-specific antibody, but notably also with the relative distribution of spike-specific cTFH subsets.^47^

Another study analysed the cellular response in samples taken from 31 COVID-19 patients, none of whom required intensive care or oxygen supplementation. A central memory phenotype (CD45RO^+^, CCR7^+^), followed by an effector memory phenotype (CD45RO+, CCR7^-^) were predominate within the spike-protein reactive CD4^+^ T cell population. An effector memory, followed by the terminal effector cells (CD45RO-, CCR7-) were the predominant phenotypes among antigen-specific CD8^+^ T cells. A significant increase in activated (CD38^+^, HLA^-^DR^+^) CD4^+^ T cells was detected among cases. Further T cell response characterisation showed CD4^+^ and CD8^+^ T cell activation in response to full-length S-protein exposure, and the M-protein response was significantly stronger (p=0.0352). A “mild” correlation between the magnitude of T-cell and humoral responses was reported (anti-RBD IgG and CD8^+^ T-cell response r=0.386 p=0.0321), whereas an interdependence was observed between the magnitude of CD8^+^ and CD4^+^ responses (r and p values not presented).^45^. Minervina and colleagues reported detection of T cell clones within both the effector and central memory subpopulations, in samples obtained from two returnees from countries with high SARS-CoV-2 transmission.^49^ Similarly, Weiskopf et al, in their study of 10 COVID-19 patients who developed ARDS, reported that peripheral SARS-CoV-2-specific CD4^+^ T-cells typically had a central memory phenotype (based on CD45RA and CCR7 expression), whereas the majority of virus-specific CD8^+^ T-cells had a CCR7- effector memory (TEM) or TEMRA phenotype.^48^ In contrast, a study of four COVID-19 positive paediatric cases with mild disease, and five uninfected controls, found no difference in the effector or central memory phenotypes of the CD8^+^ and CD4^+^ populations compared with controls.^21^

A small study conducted a phenotypic analysis of circulating SARS-CoV-2-specific T cells in samples obtained 20-47 days post positive PCR from individuals recently recovered from mild SARS-CoV-2 infection. The analysis was conducted using combination SARS-CoV-2-specific T cell detection with CyTOF. IFN-γ producing spike-specific CD4^+^ and CD8^+^ T cells were detected, suggestive of a spike-specific Th1 response, where as Th2 and Th17 lineages were not detected among spike-specific CD4^+^ T cells.^50^

Evidence of potential protective T cell mediated immunity is provided by one US-based study that measured the cellular response in rhesus monkeys (n=9 cases, n=3 controls) upon repeat challenge with pooled spike peptides. Based on IFN-γ ELISpot assays, cellular immune responses were observed in the majority of animals, with a trend toward lower responses in the lower dose groups. Intracellular cytokine staining assays demonstrated induction of both spike-specific CD4^+^ and CD8^+^ T cell responses. Post re-challenge, very limited viral RNA was observed in bronchoalveolar lavage (BAL) on day one following re-challenge in three animals, with no viral RNA detected at subsequent timepoints. In contrast, high levels of viral RNA were observed in the concurrently challenged naive animals. However, these findings to do not exclude the possibility that protection was antibody dependent rather than due to T cell immunity exclusively.^51^

## DISCUSSION

### Summary of key findings

Acutely, adult COVID-19 patients exhibit a depletion of T cells in the peripheral blood, the extent of which is positively correlated with disease severity, whereas asymptomatic patients and children tend to have preserved peripheral T cell counts. This suggests an important relationship between pathogenesis and the circulating T cell pool. It has been speculated that children may receive protection from a diverse naive T cell repertoire, with adults of increasing age at higher risk due to immunosenescence.^52^ Unfortunately, few studies have explored the relationship between T cells, age and clinical severity, with appropriate statistical adjustment. There is also emerging evidence of an important role for the over-production of cytokines – in particular IL-6 – in immunopathogenesis within COVID-19, however, drivers of these observed changes remain poorly understood.

Although less comprehensive, longer-term data suggest that T cell reductions are transient, with rapid recovery of counts within days to weeks of clinical recovery and PCR negativity. This supports the hypothesis that T cells are sequestered rather than destroyed, although the observation of similarly depleted T cell numbers in the BAL samples of severe patients indicates that T cells are not simply recruited en masse to infected tissues.^53^

In the context of well-recognised variations in COVID-19 clinical outcomes by age, ethnicity and co-morbid status, there is a striking shortage of robust evidence on demographic and clinical correlates of the cellular response to SARS-CoV-2. We identified a single study considering gender-related effects on T cells, and eight studies considering cellular responses with age (a majority of these in paediatric patients with or without adult controls). We identified no studies evaluating other potentially important determinants, including ethnicity.

Evidence characterising cellular immune responses suggest enduring T cell immunity, with phenotypic profiles consistent with helper and memory T cell functions and evidence of activity against multiple viral targets. Variation in viral targets is observed between disease severity and, based on one study, the breadth and magnitude of the T cell response were significantly higher in patients who recovered from severe compared to mild disease. Responses were also detected in individuals who experienced mild infection. However, this evidence derives from small, observational studies conducted on samples taken at varying time points from individuals, with selection criteria rarely described. The longevity of this T cell immunity and the degree of protection it provides remains unknown.

#### Strengths and limitations of the study

This study is the first to systematically evaluate and critically appraise the published literature on the T cell immune response to SARS-CoV-2, more than eight months since it has emerged. Formative reviews of evidence on the immune response are narrative with few exceptions, or focus on specific aspects.^16,54^ Our review is broader in scope and comprehensiveness.

Limitations arise from the methodology applied, and from the nature of the underlying evidence. First, while the search strategy was broad in choice of keywords and inclusion of pre-print publications, it is possible that some results were missed, particularly on pre-print servers for which structured searches are more challenging. Additional limitations arise from the nature of the underlying evidence base on which this review draws. Variations in reporting practice present major challenges for critical appraisal and weighting of evidence. For example, narrative reviews – popular in this field – have limited methods reporting. Further difficulty is introduced through the varying treatment protocols employed, clinical severity and case definitions, and varying methods adopted for T cell counts, functionality, phenotypes, and assay validation. These considerations are critical to study T cell immunity to SARS-CoV-2 as the assays are evolving and yet to be formally validated and standardised. Thirdly, many of the studies have important methodological limitations, notably; small sample sizes, minimal reporting on participation selection and reasons for follow-up, non-blinding, and widespread lack of statistical analysis that control for confounders.

#### Policy and practice implications

Many unanswered questions remain, such as the durability of and protection afforded by virus-specific T cell responses, and their relative importance in protection from reinfection compared with antibodies. More data is also needed on correlates of T cell responses and the potential of cross-reactive cellular immunity.

An important application of findings from cellular response studies will be towards the development, evaluation, and implementation of SARS-CoV-2 vaccines. In parallel with emerging data from COVID-19 patients, vaccine developers have begun to report on cellular immunogenicity from early phase evaluations, though this is notably lacking from the pre-print Phase 2 trial report of Sinovac’s inactivated vaccine.^55^ Other candidates including Moderna’s mRNA-1271, Oxford University’s ChAdOx1 nCoV-19, and CanSino’s Ad5 vaccines, have demonstrated T cell responses against S-proteins. Responses for the CanSino candidate were limited, however, by likely pre-existing immunity to the human adenovirus 5 vector. Furthermore, unlike for the second dose of Moderna’s mRNA vaccine, cellular responses were not boosted after a second dose of Oxford’s chimpanzee adenovirus-vectored vaccine.^56–58^ While there is an understandable demand for candidates able to provide protection with a single dose, vaccines which allow boosting of cellular responses may better mimic natural immunity against endemic coronaviruses. It is also worth considering whether spike-focused platforms will be able to fully harness the cellular response potential, or whether traditional inactivated whole-virus and novel virus-like particle platforms, which provide a fuller range of epitopes, will be necessary to build durable, protective immunity across populations. It will be important to evaluate vaccine efficacy in groups with high prevalence of previous exposure or infection, such as health and care staff, who will be a priority group following licensure. In addition to antibody testing, baseline assessments of virus-specific T cell reactivity are likely to be highly useful for this purpose.

Current estimates of population immunity rely solely on seroprevalence studies, however in the context of evidence for cellular responses in seronegative exposed individuals, and the apparent waning of antibody responses over time, current surveillance methods are likely to be underestimating both exposure and immunity. A more developed understanding of the role of T cells in long-term protection will be helpful to policy makers in terms of modelling where population-level immunity lies and informing long-term surveillance and immunisation strategies. However, by contrast with antibody testing – a mainstay of immune surveillance for many communicable diseases – existing T cell assays are difficult to standardise and hard to scale, therefore unlikely to be deliverable at population level within the timeframe of the SARS-CoV-2 pandemic. In the short-term, emphasis may need to be placed on determining the utility of T cell assays to guide clinical and public health actions at the individual level, particularly in patients with immunosuppression, or those at the extremes of age. In parallel, adequately powered and controlled studies providing deep immunophenotyping of T cells, B cells, and comprehensive characterisation of immune responses in mild or asymptomatic cases, and in vaccine recipients, will yield insights about the interdependence and relative importance of cellular and humoral responses. Over the long-term, development of scalable T cell assays may help to strengthen population immune surveillance systems.

#### Conclusions

A complex picture is emerging concerning the cellular immune response to SARS-CoV-2 infection, including the interplay between compartments of the immune systems, and the balance between protective versus pathological responses. Inferences are limited by methodological limitations within studies, and heterogeneity between studies. Evaluation of cellular responses at scale is currently infeasible and the benefits as yet unclear. Findings from targeted testing may carry important clinical and policy implications for public health interventions within at-risk sub-groups, for understanding mechanisms of vaccine efficacy, and for informing long-term population immunisation and surveillance strategies.

## Data Availability

This was a systematic review based on analysis of openly published secondary data.

## LIST OF ABBREVIATIONS

ARDS: Acute respiratory distress syndrome
COVID-19: Coronavirus disease (2019)
CRS: Cytokine release syndrome
ELISpot: Enzyme-linked immune absorbent spot (assay)
HCoV: Human Coronavirus (HKU1, 229E, OC43, NL63)
IFN-γ: Interferon gamma
M: Membrane (protein)
MEDLINE: Medical Literature Analysis and Retrieval System Online
MERS: Middle East Respiratory Syndrome
MetaQAT: Meta Quality Appraisal Tool
NP: Nucleocapsid protein
NSP7; NSP13: Non-structural protein 7; non-structural protein 13
PBMC: Peripheral blood mononuclear cells
PCR; RT-PCR: Polymerase chain reaction; reverse transcription polymerase chain reaction
PRISMA: Preferred Reporting Items for Systematic Reviews and Meta-Analyses
SARS-CoV-1: Severe Acute Respiratory Syndrome Coronavirus-1
SARS-CoV-2: Severe Acute Respiratory Syndrome Coronavirus-2
RBD: Receptor binding domain
RNA: Ribonucleic acid
TEMRA: T effector memory cells re-expressing CD45RA
TFH; cTFH: T follicular helper cells; circulating T follicular helper cells
Th: T helper cells
WHO: World Health Organization

## Author contributions (CRediT author statement)

MS – investigation, writing – original draft, writing – review and editing

MCIvS – conceptualisation, investigation, methodology, writing – original draft, writing – review and editing

NP – investigation, writing – original draft, writing – review and editing

DE – conceptualisation, investigation, project administration, writing – original draft, writing – review and editing

CH – investigation, writing – original draft, writing – review and editing

DL – investigation, writing – review and editing

SR – investigation, writing – review and editing

SVW – investigation, writing – review and editing

WHB – validation, writing – review and editing

PK – conceptualisation, validation, writing – review and editing

JM – validation, writing – review and editing

AMS – validation, writing – review and editing

GA – conceptualisation, supervision, validation, writing – review and editing

SJP – conceptualisation, supervision, validation, writing – review and editing

SAI – conceptualisation, investigation, methodology, project administration, writing – original draft, writing – review and editing

## Acknowledgements

We thank Professor Mike Ferguson from the School of Life Sciences, University of Dundee, for comments on the research questions and initial outputs from this work; and Professor Mark Petticrew from the Faculty of Public Health and Policy, London School of Hygiene and Tropical Medicine for advice on methodological aspects of this study.

## Ethics

This was a systematic review based on analysis of openly published secondary data. No ethical approval was required.

